# Reduced left ventricular function on cardiac MRI of SLE patients correlates with measures of disease activity and inflammation

**DOI:** 10.1101/2023.08.24.23294127

**Authors:** Audrey M. Hagiwara, Erica Montano, Gantseg Tumurkhuu, Moumita Bose, Marianne Bernardo, Daniel S. Berman, Galen Cook Wiens, Michael D. Nelson, Daniel Wallace, Janet Wei, Mariko Ishimori, C. Noel Bairey Merz, Caroline Jefferies

**Author notes:** Corresponding Author: Caroline Jefferies, PhD, Kao Autoimmunity Institute, 121 N San Vincente Blvd., Los Angeles, California, 90211. The authors have no disclosures to report.

## Abstract

**Background:** Women with SLE have an elevated risk of cardiovascular disease. Many women with SLE frequently report chest pain in the absence of obstructive coronary artery disease (CAD) due to coronary microvascular dysfunction (CMD), a form of ischemia with no obstructive CAD. Echocardiographic studies have shown that SLE patients have reduced left ventricular (LV) function, which may also correlate with higher SLE disease activity scores. As such, we used cardiac magnetic resonance imaging (cMRI) to investigate the relationship between SLE, related inflammatory biomarkers, and cardiac function in female SLE patients.

**Methods:** We performed stress cMRI in women with SLE and chest pain with no obstructive CAD (n=13, all met ACR 1997 criteria,) and reference controls (n=22) using our published protocol. We evaluated LV function, tissue characterization (T1 mapping, ECV), and delayed enhancement, using CV142 software (Circle Cardiovascular Imaging Inc, Calgary, AB, Canada). Myocardial perfusion reserve index (MPRI) was calculated using our published protocol. SLEDAI and SLICC Damage Index (DI) were calculated per validated criteria. Serum samples were analyzed for inflammatory markers and autoantibodies. Wilcoxon rank-sum test was performed on clinical values with CMD and no CMD SLE subjects, and on cMRI values with all SLE subjects and controls. Correlation analysis was done on clinical values, and cMRI values on all SLE subjects.

**Results:** Overall, 40% of SLE subjects had MPRI values < 1.84, consistent with CMD. Compared to controls, SLE subjects had significantly lower LVEF, and higher LVESVi and LVMi. Corresponding to this, radial, longitudinal, and circumferential strain were significantly lower in the SLE subjects. In correlation analysis of serum inflammatory biomarkers to cMRI values in the SLE subjects, SLICC DI was related to worse cardiac function (lower radial, circumferential and longitudinal strain) and higher T1 time. Additionally, fasting insulin and ESR were negatively correlated with LVMi. Fasting insulin also negatively correlated with ECV. CRP had a positive association with LVESV index and CI and a negative association with longitudinal strain.

**Conclusions:** Among women with SLE with chest pain and no obstructive CAD, 40% have CMD. While evaluations of known inflammatory markers (such as CRP and ESR) predictably correlated with decreased cardiac function, our study found that decreased fasting insulin levels as a novel marker of diminished LV function. In addition, low insulin levels were observed to correlate with increased LVMi and ECV, suggesting a cardioprotective effect of insulin in SLE patients. We also noted that SLICC DI, an assessment of SLE damage, correlates with cardiac dysfunction in SLE. Our findings underline the potential of non-invasive cMRI as a tool for monitoring cardiovascular function in SLE, particularly in patients with high SLICC DI, ESR and CRP and low fasting insulin levels.

## Introduction

Systemic lupus erythematosus (SLE) is a sexually dimorphic autoimmune disease, with approximately 90% of patients being women. Chest pain is a common symptom in patients with SLE [1], with cardiovascular disease (CVD) being a significant contributor to morbidity and mortality in SLE. Compared to the general population, cardiovascular mortality is 2.7-fold higher in SLE patients [2]. The risk of CVD is significantly elevated in women with SLE, particularly in the 35 to 44 year age group who have a 50-fold increased risk of myocardial infarction (MI) compared to age-matched healthy individuals [3]. The origin of chest pain in SLE can be attributed to multiple causes, including pericardial disease, myocarditis and coronary artery disease (CAD). However, coronary angiography is frequently normal, indicating causes other than CAD driving anginal chest pain [4].

With the advancement in non-invasive diagnostic imaging, it is now understood that many SLE patients have ischemia with no obstructive CAD (INOCA), which is associated with elevated risk for CVD events [5-8]. Studies using cardiac magnetic resonance imaging (cMRI) have demonstrated a high prevalence of INOCA amongst SLE patients, which is predominantly attributable to coronary microvascular dysfunction (CMD) [9-11]. Now a routine tool within cardiac diagnostics, cMRI measures determines the mass and volumes of the heart, in addition to providing structural imaging of the myocardial tissue to detect fibrosis. Stress perfusion imaging allows measurement of coronary blood flow, which permits detection of ischemia and valve disorders. The semi-quantitative myocardial perfusion index (MPRI) is both sensitive and specific for the diagnosis of CMD (a MPRI of less than 1.84 being diagnostic of CMD), whereas measures of left ventricular diastolic volumes and ejection fraction detect cardiac dysfunction [12]. CMRI MPRI is an independent predictor of major adverse cardiovascular events (MACE) in patients with INOCA [13]. In addition to cardiac positron emission tomography (PET), which is the most studied non-invasive method for diagnosing CMD, stress cMRI is a reliable tool for diagnosing CMD while also offering insight into cardiac anatomy and function [12, 14-16].

The pathogenic mechanisms underlying CVD in SLE are incompletely understood; however, inflammation, endothelial dysfunction, and autoimmunity are known contributing factors. For example, systemic inflammation results in the upregulation of endothelial adhesion molecules, increased recruitment of mononuclear cells, and the production of pro-inflammatory cytokines which propagate atherogenesis [17, 18]. Clinical measurements of inflammation in SLE include complement, erythrocyte sedimentation rate (ESR), and C-reactive protein (CRP), which are used in scoring tools to determine and follow SLE disease activity and organ damage. Activation of the complement system leads to low levels of C3 and C4 in SLE [19]. ESR and CRP are non-specific markers of inflammation that are elevated in SLE and are associated with organ inflammation and infection, respectively [20]. It is unknown whether markers of clinical inflammation in SLE correlate with cardiac function abnormalities on cMRI. As such, this study aims to correlate changes in cardiac function in SLE patients using advanced cardiac imaging with clinical, laboratory and disease activity profiles of the patients in order to better understand subclinical heart disease in SLE.

## Methods

Patients and Methods: This cross-sectional study was approved by the institutional review board at Cedars-Sinai Medical Center, and all participants gave informed consent prior to participation. Participants were recruited from the rheumatologic clinic. Inclusion criteria consisted of female SLE subjects (aged 37 to 57 years at baseline) with chest pain due to suspected angina. Exclusion criteria included documented obstructive CAD and contraindications to coronary computed tomography angiography (CCTA) or cardiac magnetic resonance imaging (cMRI).

Clinical Evaluation: Demographic data of the patients were collected including age and smoking history. Participants completed standardized angina symptom questionnaires and quality of life measures related to angina, in addition to a demographic questionnaire [21, 22]. Clinical data was collected including vital signs and body mass index. SLE disease activity was collected using the SLE disease activity index (SLEDAI) [23] and the systemic lupus international collaborating clinics (SLICC) Damage Index (DI) [24]. In addition, serum samples were taken and evaluated for complete blood count, creatinine, protein, fasting blood sugar, fasting insulin, inflammatory markers (erythrocyte sedimentation rate (ESR), C-reactive protein (CRP), Complements (C3, C4)), and antibodies (anti-antinuclear antibody (ANA), anti-double stranded DNA (DNA), ribonucleoprotein (RNP), anti-Smith (Sm), SSA (Ro), SSB (La), and anti-topoisomerase I (Scl)). Obstructive CAD was excluded by noninvasive CCTA.

Cardiac Magnetic Resonance Imaging: Using a standardized cardiac magnetic resonance imaging (cMRI) protocol [25], participants underwent stress cMRI at 3.0-Tesla (Siemens Vida or Biograph, Erlangen Germany) using ECG-gating/ phased array coil, with 0.05 mmol/kg gadolinium first-pass perfusion three slice stress (adenosine 140 μg/kg-1/min-1 or regadenoson 0.4 mg) followed by rest, function, myocardial characterization (T1 and T2 mapping), and late gadolinium enhancement. T1 mapping was acquired using the modified Look-Locker inversion recovery (MOLLI) method. Caffeine was withdrawn for 24 hours prior to stress testing. Beta blocker, calcium channel blocker and nitrate medications were with-held for 24-48 hours prior to stress testing, at physician discretion. Blood was collected for hematocrit measurement on the day of imaging for calculating extracellular volume (ECV) fraction.

Cardiac MRI Analysis: cMRI studies were analyzed for LV function, strain, volumes, cardiac output, mass, T1 and T2 mapping, using CVI42 software (Circle Cardiovascular Imaging Inc, Calgary, AB, Canada), by the Cedars-Sinai Medical Center cMRI Core Laboratory. Myocardial perfusion reserve index (MPRI) was calculated as previously published [12]. LV mass and end-systolic and end-diastolic volumes were indexed to the body surface area (BSA). Native T1, ECV, and T2 times were measured in the midseptal region of the midventricular slice. Abnormal cMRI demonstrating coronary microvascular dysfunction was defined as a MPRI less than 1.84.

Statistical Analysis: Measurements are expressed as a mean and standard deviation, median and range for continuous variables, and counts and percentages for categorical variables. Groups were compared using t tests for SLE vs RC, with Welch’s correction for unequal variances. Wilcoxon rank-sum test was used to analyze clinical and cardiac function data between patients with CMD and those with no CMD. One way ANOVA was used to compare among CMD, no CMD and HC. When significant, a Bonferroni correction was used for post hoc pair-wise tests. Spearman correlations were used to assess association between CMRI features and lab results. Effect size was calculated as Cohen’s d with 95% confidence intervals for comparing between CMD and no CMD on CMRI features. A significance level of 0.05 was used for all hypothesis tests.

## Results

Female SLE patient baseline characteristics are summarized in **Table 1**. The average age of the patients (n=13) recruited was 47, with SLEDAI scores ranging from inactive (0-3) to moderately active (4-6). Vital signs and blood count were all normal and clinical labs were consistent with a diagnosis of SLE, with serum creatinine and C reactive protein (CRP) being elevated compared to reference controls, whilst complement proteins C3 and C4 were reduced. Interestingly, while systolic blood pressure, body mass index (BMI) and heart rate were all similar between reference controls and SLE patients, diastolic blood pressure (DBP) was raised in SLE patients compared to reference controls (78.4+9.3 vs. 59+11 mmHg, respectively).

**Table 1:** Baseline SLE Subject Characteristics. Results are presented as mean ± SD or median (min-max). Normal reference ranges are given in column 2 in square brackets.

| | Mean ± S.D. | Median (min-max) |
| --- | --- | --- |
| **Age** | 45 ± 5 years | 44 (36-51) years |
| **Ethnicity** |  |  |
| Hispanic | 3 |  |
| Non-Hispanic | 10 |  |
| **SLEDAI** | 2.5 ± 2.5 | 2 (0-6) |
| **Clinical Labs** |  |  |
| Fasting Blood Sugar | 77.7 ± 7.9 mg/dl [100-125 mg/dl] | 79 (65-90) mg/dl |
| Fasting Insulin | 9.6 ± 13.4 mIU/ml [2-20 mIU/ml] | 5.2 (1.4-41.5) mIU/ml |
| Serum Creatinine | 1.26 ± 2.2 mg/dL [0.59-1.04 mg/dL] | 0.72 (0.55-5.4) mg/dl |
| Serum Protein | 7.17 ± 1.04 g/dL [6-8 g/dL] | 6.9 (6-10.1) g/dl |
| C3 | 93.5 ± 4.5 mg/dL [80-160 mg/dL] | 92 (70-153) mg/dl |
| C4 | 21.75 ± 7.5 mg/dL [15-45 mg/dL] | 22 (10-44) mg/dl |
| ESR | 16.8 ± 20.38 mm/hr [0-29mm/hr] | 12 (1-73) mm/hr |
| CRP | 1.84 ± 1.85 mg/dL [<1mg/dL] | 0.8 (0.2-5.4) mg/dl |
| **Vital Signs** |  |  |
| Systolic (mmHg) | 115.8 ± 13.4 | 111.5 (99-143) |
| Diastolic (mmHg) | 76.8 ± 11.3 | 76.5 (58.5-97) |
| Pulse | 69.8 ± 16.8 bpm | 65 (49-98) bpm |
| Respiration | 16.9 ± 1.1 [12-20 BPM] | 17 (15-18) BPM |
| BMI | 25.3 ± 5.3 | 27.1 (18-33) |
| **Blood count** |  |  |
| Hemoglobin | 12.6 ± 1.3 g/dL [11.6-15 g/dL] | 12.6 (10-14.9 g/dL) |
| WBC | 4.6 ± 1.5 $10^9$/L [4-10 $10^9$/L] | 4.35 (2.27-7.24 $10^9$/L) |
| Platelets | 250 ± 41 $10^9$/L [157-371 $10^9$/L] | 249 (186-311 $10^9$/L) |

CMRI results are shown in **Table 2**, comparing cardiac function of SLE patients with a reference control group from the same center (n=22, all female). Of note, the age range of patients and reference controls was similar (47+6 vs. 49+6 years, respectively). LV ejection fraction was significantly decreased in SLE patients compared to reference controls (59±7% compared to 64±5%, p=0.0291). LV mass index (LVMi) was also significantly increased compared to reference controls (47.31±9.3 ml/m^2^ compared to 41.34.5±4.3 ml/m^2^, p= 0.0461). In addition, radial, circumferential and longitudinal strain were reduced significantly in SLE patients compared to reference controls. Of the SLE patients, none had evidence of segmental perfusion defects, consistent with absence of obstructive CAD. One subject had LV ejection fraction <50% with normal perfusion and no late gadolinium enhancement, consistent with nonischemic cardiomyopathy. An additional 2/13 (15%) had evidence of late gadolinium enhancement; one with extensive patchy scar in the absence of elevated T2 and normal wall motion, consistent with prior myocarditis, and another with small basal midmyocardial scar of uncertain significance. Mean T2 relaxation time was 43±4 ms (range 36-50 ms).

**Table 2:**
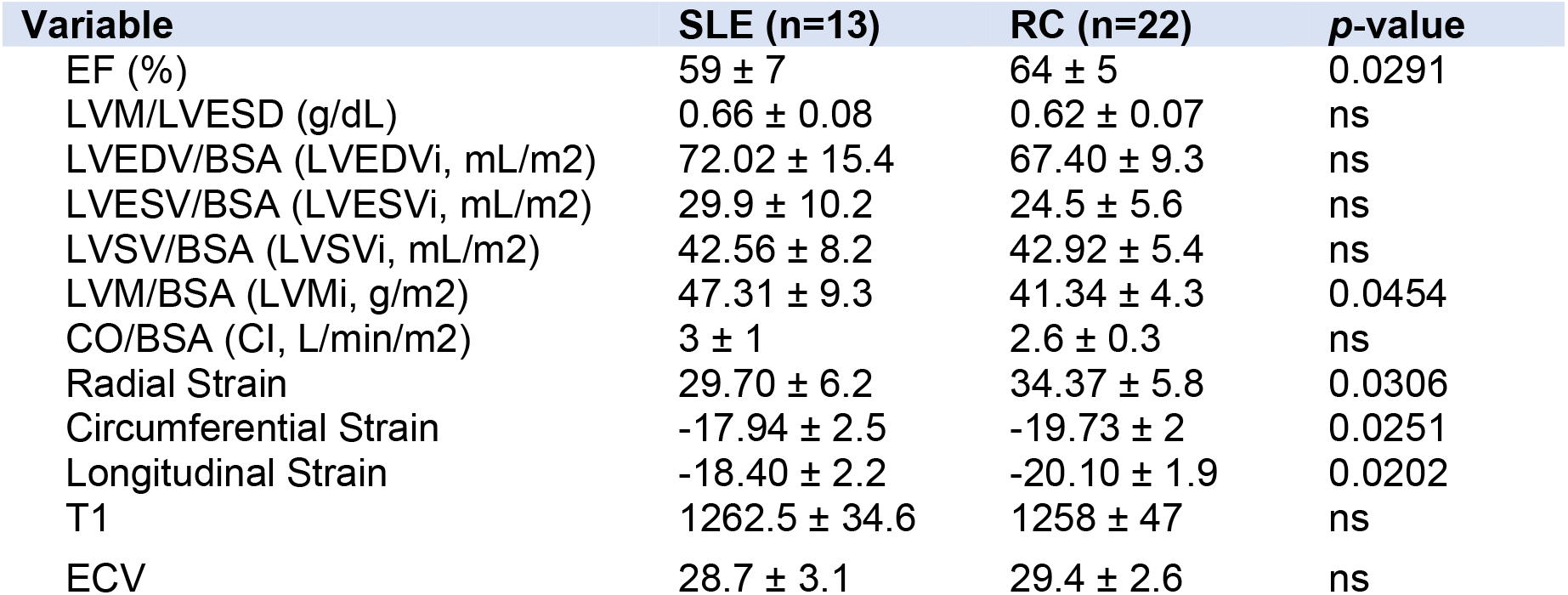
Comparison of cardiac function between SLE patients and reference controls based on values obtained on cardiac MRI. Results are shown as mean±S.D. and significance was determined using student t test with or without Welch’s correction as appropriate. LVESV: left ventricular end-systolic volume; LVEDV: left ventricular end-diastolic volume; LVSV: left ventricular systolic volume; EF: ejection fraction; LVM: left ventricular mass; BSA: body surface area; ECV: extracellular volume. ns represents a *p*-value > 0.05.

A Spearman correlation matrix was constructed to determine potential relationships between cardiac function measurements and clinical laboratory findings. Associations are shown in **Table 3**, with positive associations in green and negative associations in orange. We observed that the SLICC disease index (DI) score, which measures accumulated organ damage since onset of SLE, negatively correlated with LVEF, measures of strain but positively correlated with native T1 times. Fasting insulin levels and ESR showed a strong negative correlation with LVMi (Spearman r=-0.664 and -0.604, respectively) and fasting insulin levels also negatively correlated with extracellular volume (ECV) (r=-0.664). C reactive protein (CRP), a marker of inflammation, showed a moderate-strong positive association with LVESVi and cardiac index and a similar trend towards a moderate negative association with longitudinal strain.

**Table 3:**
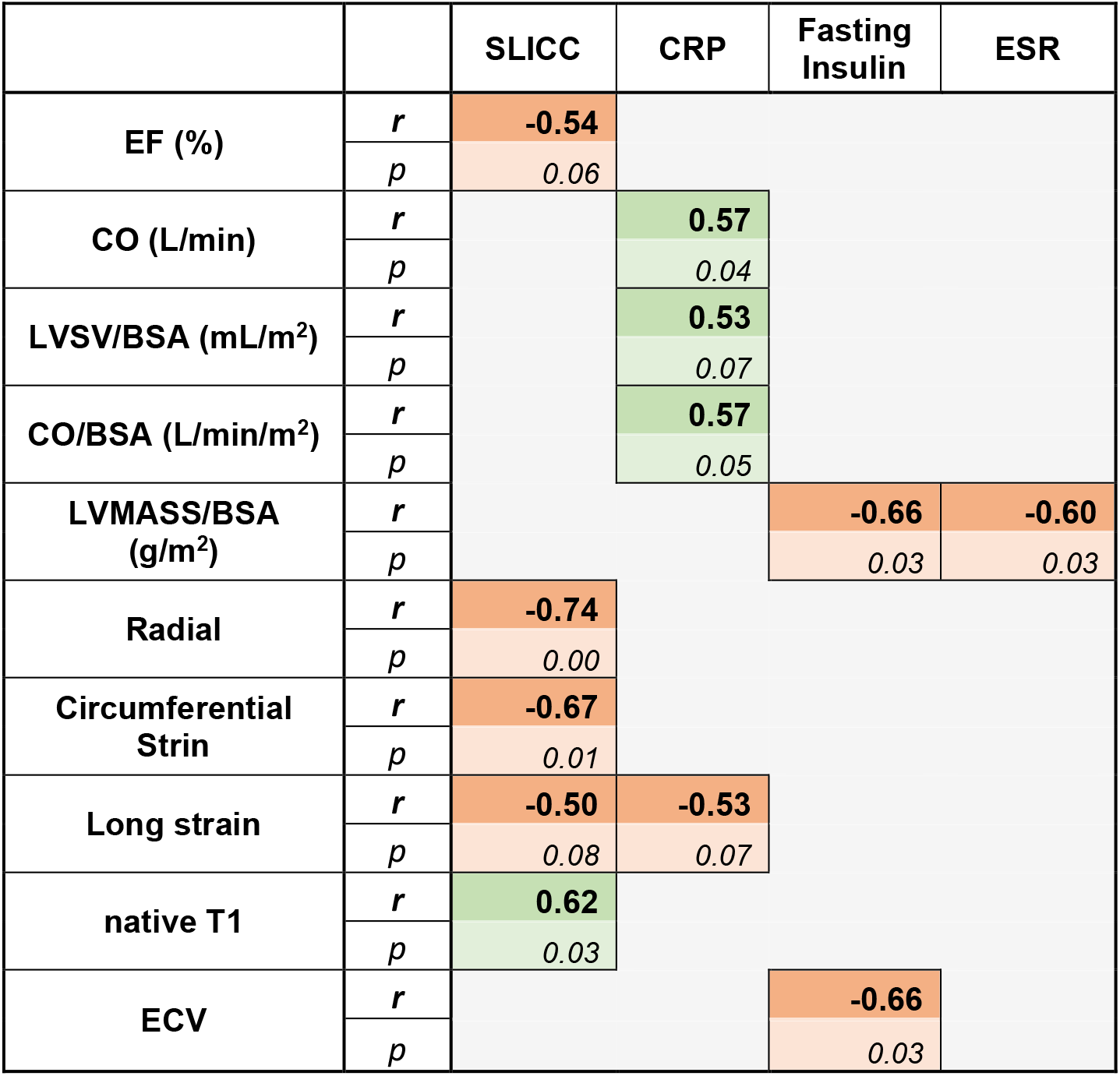
Analysis of cardiac function versus clinical values from patients in the study. A correlation matrix was constructed to determine potential relationships between cardiac function measurements on cMRI and clinical laboratory findings. Associations are shown and are represented as positive associations in the green boxes and negative associations in the orange boxes. The Spearman *r* coefficient and *p* value is shown in each case. LVSV: left ventricular systolic volume; EF: ejection fraction; CO: Cardiac output; LVM: left ventricular mass; BSA: body surface area; ECV: extracellular volume.

We next analyzed the data based on whether SLE patients had CMD. Similar to previous work, we found that 5/13 (39%) of SLE subjects had CMD. No differences were observed between SLE patients with or without CMD when vital signs or clinical labs were compared **(Table 4)**. When SLE cardiac function was assessed using Wilcoxon rank-sum test, we observed that left ventricular function was significantly different between patients with CMD or no CMD **(Table 5)**. However no difference in EF or myocardial strain was observed between patients with or without CMD. However, we noted that patients without CMD had higher LVMi compared with either patients with CMD or reference controls **(Table 5)**, prompting us to assess the relative contribution of CMD versus no CMD to the overall differences in cardiac function between reference controls and SLE patients – ie ejection fraction, myocardial strain and LVMi. As shown in **Figure 1**, we observe that cardiac function was more similar between patients with CMD and reference controls, indicating that patients without CMD had a larger degree of contribution to the overall effect size noted previously. **Figure 2** demonstrates how the patient group without CMD contributed to the overall differences observed. Overall, SLE patients without CMD (blue) contributed stronger to the effect seen between reference control cardiac MRI function than did patients with CMD (black).

**Table 4:** Baseline SLE Subject Characteristics grouped into patients with CMD (CMD) or without CMD (no CMD). Results are presented as mean±S.D [min-max values]. Data was analyzed by unpaired student’s t test, with Welch’s correction as appropriate. In all cases, no significant difference between the two groups was observed.

| Mean ±SD (range) | CMD (n=5) | No CMD (n=8) |
| --- | --- | --- |
| **Age** | 43 (37-51) years | 48 (43-57) years |
| **SLEDAI** | 2.8 ± 3 [0-6] | 1.4 ± 2 [0-6] |
| **SLICC** | 0.8 ± 0.8 [0-2] | 0.4 ± 1 [0-3] |
| **Clinical Labs** |  |  |
| Fasting Blood Sugar | 80.2 ± 6.5 mg/dl [72-90] | 75.5 ± 10.1 mg/dl [65-91] |
| Fasting Insulin | 14.8 ± 15.4 mIU/ml [2-42] | 4.5 ± 2.5 mIU/ml [1.4-7.6] |
| Serum Creatinine | 0.67 ± 0.12 mg/dL [0.55-0.84] | 0.72 ± 0.03 mg/dl [0.66-0.76] |
| Serum Protein | 7.58 ± 1.47 g/dL [6.4-10.1] | 7.09 ± 0.40 g/dl [6.8-7.9] |
| C3 | 124 ± 34 mg/dL [81-153] | 108 ± 26 mg/dl [77-156] |
| C4 | 20 ± 11 mg/dL [10-32] | 29 ± 12 mg/dl [22-51] |
| ESR | 32 ± 24 mm/hr [16-73] | 14 ± 16 mm/hr [1-47] |
| CRP | 2.6 ± 2.2 mg/dL [0.4-5.4] | 3.6 ± 6.0 mg/dl [0.2-17.9] |
| Anti-ANA | 424 ± 299 [50-640] | 440 ± 277 [80-640] |
| Anti-DNA | <10 | <10 |
| RNP Antibody | 68 ± 70.4 [2-156] | 26.3 ± 46.8 [2-133] |
| Anti-Sm | 17.2 ± 20.6 [2-52] | 11.9 ± 18.6 [1-51] |
| SSA (Ro) | 42.8 ± 55 [2-104] | 10.4 ± 19.2 [1-56] |
| SSB (La) | 22.8 ± 45.5[2-104] | 5.5 ± 8.3 [1-25] |
| Anti-SCL | 4.2 ± 3.3 [2-10] | 1 ± 25 [1-25] |
| **Vital Signs** |  |  |
| Systolic (mmHg) | 116.8 ± 7.4 [109-128] | 119.4 ± 14 [104-127] |
| Diastolic (mmHg) | 78 ± 8.7 [70-92] | 78.6 ± 10 [66-87] |
| Pulse (bpm) | 79 ± 13 [72-90] | 65 ± 16 [49-89] |
| BMI (kg/m<sup>2</sup>) | 24.2 ± 4.9 [18-28] | 27.5 ± 5.2 [19-33] |
| **Blood count** |  |  |
| Hemoglobin | 13.0 ± 1.6 g/dL [11.2-14.9] | 12.6 ± 1.0 g/dL [11.1-13.9] |
| WBC | 5.0 ± 1.37 10<sup>9</sup>/L [3.4-6.8] | 4.4 ± 1.58 10<sup>9</sup>/L [3.1-7.2] |
| Platelets | 256 ± 41 10<sup>9</sup>/L [157-371] | 224 ± 46 10<sup>9</sup>/L [173-305] |

**Table 5:** Comparison of cardiac function in SLE patients with CMD compared to SLE patients without on cMRI. Wilcoxan rank-summed test was used to determine potential differences in cardiac function between patients with CMD (Y, N=5) versus those without (N, N=8). LVESV: left ventricular end-systolic volume; LVEDV: left ventricular end-diastolic volume; LVSV: left ventricular systolic volume; LVM: left ventricular mass; BSA: body surface area.

|  | **N**<br>**(N=8)** | **Y**<br>**(N=5)** | **Total**<br>**(N=13)** | **p value** |
| --- | --- | --- | --- | --- |
| **LVEDV/BSA (mL/m2)** |  |  |  | 0.0068 |
| Mean (SD) | 79.8 (13.2) | 59.5 (9.5) | 72.0 (15.4) |  |
| Median | 76.8 | 60.3 | 70.7 |  |
| Q1, Q3 | 70.8, 84.8 | 56.4, 65.2 | 65.2, 80.7 |  |
| Range | (66.5-107.4) | (45.4-70.4) | (45.4-107.4) |  |
| **LVESV/BSA (mL/m2)** |  |  |  | 0.1243 |
| Mean (SD) | 33.4 (10.7) | 24.4 (6.9) | 29.9 (10.2) |  |
| Median | 29.3 | 23.3 | 27.4 |  |
| Q1, Q3 | 26.4, 42.6 | 22.0, 27.1 | 23.3, 34.1 |  |
| Range | (20.6-50.1) | (15.3-34.1) | (15.3-50.1) |  |
| **LVM/BSA (g/m2)** |  |  |  | 0.0233 |
| Mean (SD) | 52.5 (6.9) | 39.0 (5.9) | 47.3 (9.3) |  |
| Median | 53.4 | 35.7 | 45.6 |  |
| Q1, Q3 | 46.7, 58.1 | 35.3, 45.1 | 42.8, 55.6 |  |
| Range | (42.8-61.1) | (33.2-45.6) | (33.2-61.1) |  |
| **LVSV/BSA (mL/m2)** |  |  |  | 0.0481 |
| Mean (SD) | 46.4 (7.3) | 36.4 (5.5) | 42.6 (8.2) |  |
| Median | 44.6 | 36.3 | 42.4 |  |
| Q1, Q3 | 41.5, 52.8 | 33.0, 38.2 | 36.3, 45.5 |  |
| Range | (36.1-57.4) | (30.0-44.4) | (30.0-57.4) |  |

**Figure 1.**
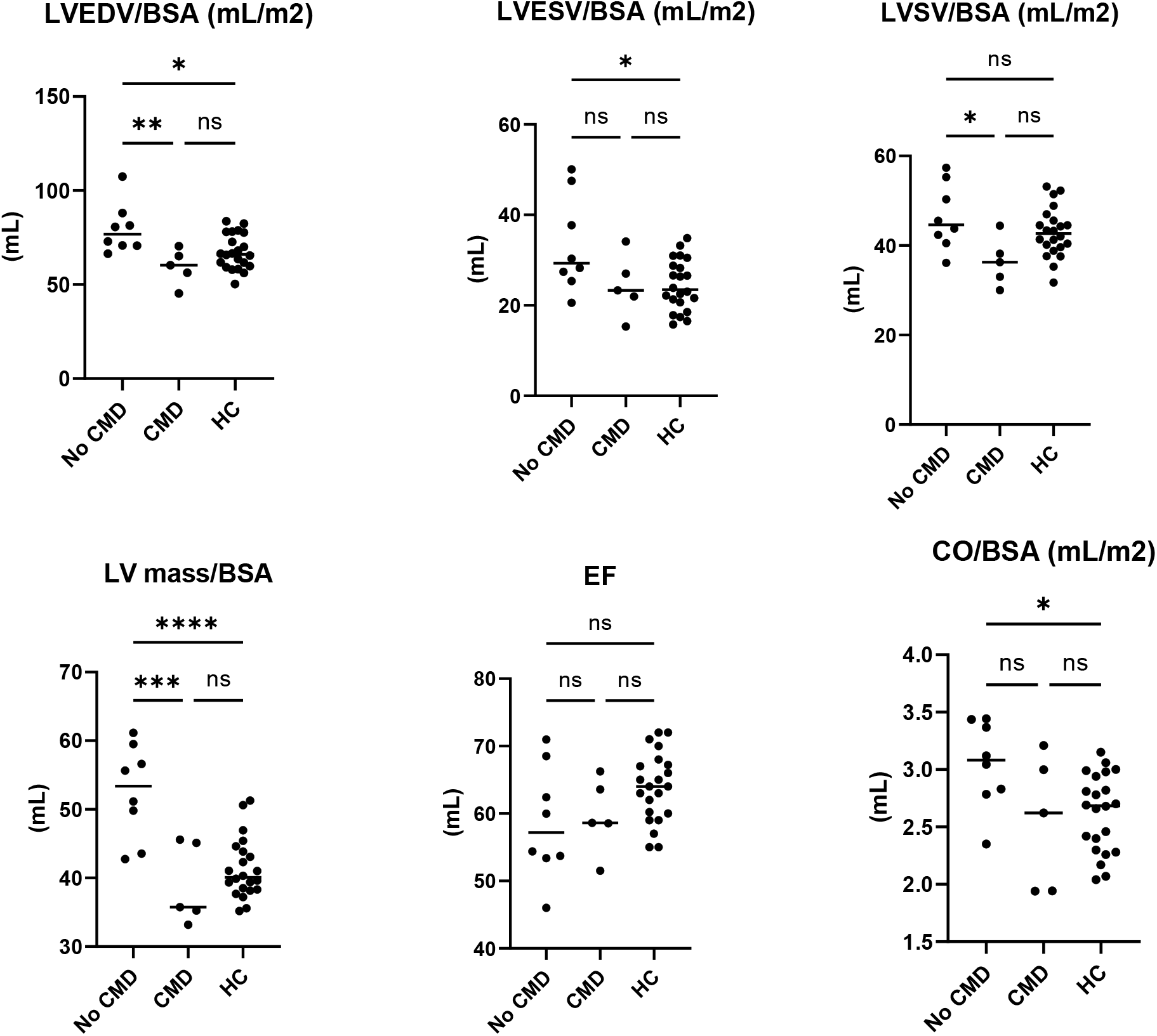
Comparison of cardiac function in SLE patients with CMD or no CMD compared to reference controls. Statistical analysis was performed using One-Way ANOVA and Tukey’s multiple comparison test. ****p<0.0001, ***p<0.001, \*\**p* <0.01, *p<0.05, ns= not significant.

**Figure 2:**
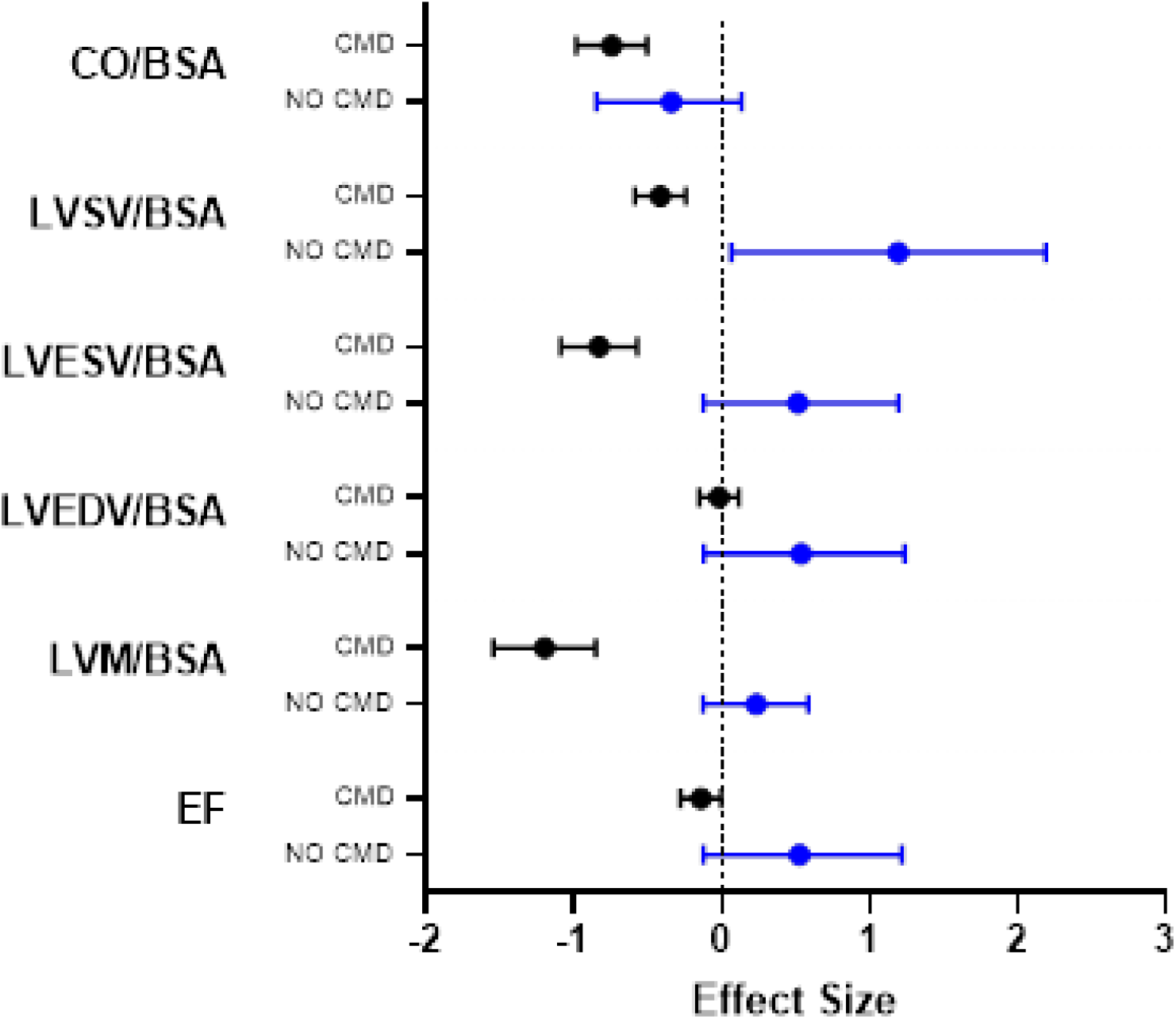
Patients without CMD (no CMD) show a stronger contribution to differences in cardiac function between healthy controls and SLE patients. Patients with no CMD (blue) on MPRI show a stronger contribution to overall effect of SLE on cardiac function than do patients with CMD (black bars) when compared with reference controls. Data is represented as Cohen’s *d* ± C.I.

## Discussion

Vascular changes are well known to occur as a result of systemic inflammation, such as experienced by patients with SLE. The present study indicates that SLE patients have decreased LV ejection fraction, together with decreased myocardial strain and increased LV mass (adjusted for body surface area (LVMi)). Importantly, inflammatory markers such as CRP and ESR and the SLICC DI correlated with changes in cardiac function on cMRI, indicating that chronic inflammation may be driving these changes. SLICC DI positively correlated with native T1 time, indicating that increased damage may be linked to increased risk of cardiac fibrosis. Interestingly, decreased fasting insulin levels were moderately associated with both increased LV mass and extracellular volume fraction (ECV). These results are intriguing as a recent study identified low fasting insulin levels as an independent predictor of all-cause mortality and cardiovascular mortality in patients with acute decompensated heart failure without diabetes mellitus [26]. Similarly, insulin is known to play a role in adaption to myocardial stress [27], and has also been identified as having anti-inflammatory effects in rodents with an acute phase response after burn injury, reducing levels of pro-inflammatory cytokines [28]. Insulin also affects fatty acid lipolysis, leading to reduced plasma fatty acid levels [29]. Thus low insulin levels in SLE patients may be an important prognostic factor in cardiac dysfunction.

Previous findings have reported alterations in LV function in patients with SLE, in part associated with traditional risk factors of age, obesity, hypertension and diabetes [30]. Our study is in agreement with these previous studies, reporting increased LVMi, decreased LV ejection fraction and decreased myocardial strain. These changes reflect global LV remodeling as described in earlier studies and are in indicative of LV dysfunction in our SLE cohort [30-33].

Impairment in longitudinal strain has been suggested as an index for evaluating ventricular function. Recent studies using speckle tracking echocardiography (STE) have reported that SLE patients show impairment in cardiac strain reflecting cardiac dysfunction [34-36]. Nikdoust *et al* reported that SLE patients with no reported cardiac symptoms had significant decreases in longitudinal strain [34], whilst Taha *et al* showed that SLE patients with active disease had impairment in longitudinal, circumferential and radial strain on STE [35]. Meanwhile Zhong et al demonstrated that disease activity and markers of inflammation (specifically CRP, ESR and C3/C4) correlated with decreased speckle strain parameters, indicating subclinical myocardial injury [36]. Both ESR and CRP are non-specific markers of inflammation, being increased in malignancy, infection and autoimmune disease [37]. In SLE, both markers tend to increase with disease exacerbations, making them potentially useful biomarkers of disease activity [20]. CRP levels have previously been associated with cardiac abnormalities in SLE patients on cMRI [6, 38]. In line with these studies we have observed impaired cardiac strain on cMRI, with radial strain decreased by approximately 20% from reference controls from the same center and circumferential and longitudinal strain approximately 10% changed. In addition, cardiac strain and LV function (EF, LVSV and LVESV) show a moderate to strong correlation with SLICC DI, as a measure of damage accrual in SLE, in addition to markers of systemic inflammation and disease activity such as CRP and ESR. SLICC and CRP levels also correlate with changes in native T1 time and ECV, both of which are indicative of cardiac fibrosis, a strong predictor of subclinical LV dysfunction. Interestingly, previous studies have shown that higher SLICC DI scores have been associated with macrovascular effects, with studies demonstrating SLE subjects with higher SLICC DI scores to have higher risk of developing MACE [39] and increased plaque development [40, 41]. However, in our study, we have demonstrated that SLICC DI may also be a useful tool in identifying microvascular effects and subclinical atherosclerotic disease.

Although the SLE subjects with CMD were overall similar to the subjects without CMD, subjects without CMD had higher LV mass index compared to either SLE subjects with CMD or reference controls. This may have been driven by two SLE subjects who had MPRI>1.8 and the highest LV mass index and end-diastolic volume index of the group, possibly representing subclinical nonischemic SLE-related dilated cardiomyopathy phenotype. Since late gadolinium enhancement and myocardial T2 were normal in these two subjects, SLE myocarditis is an unlikely pathophysiologic mechanism for this finding. While LV mass index is predictive of adverse cardiovascular events including heart failure [42-44], a larger sample size is needed to determine whether CMD in SLE patients with chest pain is related to lower LV mass index or whether the absence of CMD suggests an alternative chest pain pathophysiology such as coronary vasospasm that may be related to higher LV mass index. Since cMRI can identify early pathophysiologic structural and functional changes prior to the onset of clinically over cardiovascular disease, cMRI has potential to enhance the understanding of SLE disease activity [45].

### Limitations

This study has several limitations that should be addressed. First, this was a single center study with a small sample size. Future prospective studies with larger populations are required. Secondly, invasive coronary function testing was not carried out to diagnose coronary vasospasm to explain the patients without CMD but with chest pain had an alternative chest pathophysiology. Further research is required to determine this. Thirdly, further research is required to understand the contribution low insulin levels play in cardiac dysfunction in SLE, such as molecular assessment of immune cell responses to insulin and whether insulin signaling contributes to reduction of inflammation in SLE immune cells.

## Conclusions

This study confirms the association between markers of inflammation in autoimmune disease such as ESR and CRP potential subclinical atherosclerotic disease in SLE patients on cMRI. It also reveals for the first time that patients with high SLICC DI may be more at risk for developing changes in left ventricular function and subclinical CVD. Similarly, low insulin levels were observed to correlate with increased LVMi and ECV, suggesting a cardioprotective effect of insulin in SLE patients Our findings underline the potential of non-invasive cMRI as a tool for monitoring cardiovascular function in SLE, particularly in patients with high SLICC DI, ESR and CRP and low fasting insulin levels.

## Data Availability

All data produced in the present study are available upon reasonable request to the authors

## References

1. Ishimori, M.L., et al., Microvascular angina: an underappreciated cause of SLE chest pain. J Rheumatol, 2013. 40(5): p. 746–7.

2. Yurkovich, M., et al., Overall and cause-specific mortality in patients with systemic lupus erythematosus: a meta-analysis of observational studies. Arthritis Care Res (Hoboken), 2014. 66(4): p. 608–16.

3. Manzi, S., et al., Age-specific incidence rates of myocardial infarction and angina in women with systemic lupus erythematosus: comparison with the Framingham Study. Am J Epidemiol, 1997. 145(5): p. 408–15.

4. Ishimori, M.L., et al., Prevalence of angina in patients with systemic lupus erythematosus. Arthritis Research & Therapy, 2012. 14(3): p. A62.

5. Manchanda, A.S., et al., Coronary Microvascular Dysfunction in Patients With Systemic Lupus Erythematosus and Chest Pain. Front Cardiovasc Med, 2022. 9: p. 867155.

6. Sandhu, V.K., et al., Five-Year Follow-Up of Coronary Microvascular Dysfunction and Coronary Artery Disease in Systemic Lupus Erythematosus: Results From a Community-Based Lupus Cohort. Arthritis Care Res (Hoboken), 2020. 72(7): p. 882–887.

7. Bairey Merz, C.N., et al., Ischemia and No Obstructive Coronary Artery Disease (INOCA): Developing Evidence-Based Therapies and Research Agenda for the Next Decade. Circulation, 2017. 135(11): p. 1075–1092.

8. Weber, B.N., et al., Coronary Microvascular Dysfunction in Systemic Lupus Erythematosus. J Am Heart Assoc, 2021. 10(13): p. e018555.

9. Kiani, A.N., et al., Coronary calcification in SLE: comparison with the Multi-Ethnic Study of Atherosclerosis. Rheumatology (Oxford), 2015. 54(11): p. 1976–81.

10. Gartshteyn, Y., et al., Prevalence of coronary artery calcification in young patients with SLE of predominantly Hispanic and African-American descent. Lupus Sci Med, 2019. 6(1): p. e000330.

11. Ishimori, M.L., et al., Myocardial ischemia in the absence of obstructive coronary artery disease in systemic lupus erythematosus. JACC Cardiovasc Imaging, 2011. 4(1): p. 27–33.

12. Thomson, L.E., et al., Cardiac magnetic resonance myocardial perfusion reserve index is reduced in women with coronary microvascular dysfunction. A National Heart, Lung, and Blood Institute-sponsored study from the Women’s Ischemia Syndrome Evaluation. Circ Cardiovasc Imaging, 2015. 8(4).

13. Zhou, W., et al., Long-Term Prognosis of Patients With Coronary Microvascular Disease Using Stress Perfusion Cardiac Magnetic Resonance. JACC Cardiovasc Imaging, 2021. 14(3): p. 602–611.

14. Zorach, B., et al., Quantitative cardiovascular magnetic resonance perfusion imaging identifies reduced flow reserve in microvascular coronary artery disease. J Cardiovasc Magn Reson, 2018. 20(1): p. 14.

15. Kotecha, T., et al., Automated Pixel-Wise Quantitative Myocardial Perfusion Mapping by CMR to Detect Obstructive Coronary Artery Disease and Coronary Microvascular Dysfunction: Validation Against Invasive Coronary Physiology. JACC Cardiovasc Imaging, 2019. 12(10): p. 1958–1969.

16. Marano, P., J. Wei, and C.N.B. Merz, Coronary Microvascular Dysfunction: What Clinicians and Investigators Should Know. Curr Atheroscler Rep, 2023.

17. Bartoloni, E., Y. Shoenfeld, and R. Gerli, Inflammatory and autoimmune mechanisms in the induction of atherosclerotic damage in systemic rheumatic diseases: two faces of the same coin. Arthritis Care Res (Hoboken), 2011. 63(2): p. 178–83.

18. Prasad, M., et al., Cardiorheumatology: cardiac involvement in systemic rheumatic disease. Nat Rev Cardiol, 2015. 12(3): p. 168–76.

19. Weinstein, A., R.V. Alexander, and D.J. Zack, A Review of Complement Activation in SLE. Curr Rheumatol Rep, 2021. 23(3): p. 16.

20. Aringer, M., Inflammatory markers in systemic lupus erythematosus. J Autoimmun, 2020. 110: p. 102374.

21. Rose, G.A., The diagnosis of ischaemic heart pain and intermittent claudication in field surveys. Bull World Health Organ, 1962. 27(6): p. 645–58.

22. Rahman, M.A., et al., Rose Angina Questionnaire: validation with cardiologists’ diagnoses to detect coronary heart disease in Bangladesh. Indian Heart J, 2013. 65(1): p. 30–9.

23. Bombardier, C., et al., Derivation of the SLEDAI. A disease activity index for lupus patients. The Committee on Prognosis Studies in SLE. Arthritis Rheum, 1992. 35(6): p. 630–40.

24. Petri, M., et al., Derivation and validation of the Systemic Lupus International Collaborating Clinics classification criteria for systemic lupus erythematosus. Arthritis Rheum, 2012. 64(8): p. 2677–86.

25. Aldiwani, H., et al., Reduced myocardial perfusion is common among subjects with ischemia and no obstructive coronary artery disease and heart failure with preserved ejection fraction: a report from the WISE-CVD continuation study. Vessel Plus, 2022. 6.

26. Nogi, M., et al., Low Insulin Is an Independent Predictor of All-Cause and Cardiovascular Death in Acute Decompensated Heart Failure Patients Without Diabetes Mellitus. J Am Heart Assoc, 2020. 9(10): p. e015393.

27. Rahman, A., et al., Malnutrition and cachexia in heart failure. Journal of parenteral and enteral nutrition, 2016. 40(4): p. 475–486.

28. Jeschke, M.G., et al., Effect of insulin on the inflammatory and acute phase response after burn injury. Critical care medicine, 2007. 35(9): p. S519–S523.

29. Grossman, A.N., et al., Glucose-insulin-potassium revived: current status in acute coronary syndromes and the energy-depleted heart. Circulation, 2013. 127(9): p. 1040–1048.

30. Pieretti, J., et al., Systemic lupus erythematosus predicts increased left ventricular mass. Circulation, 2007. 116(4): p. 419–26.

31. Gaudron, P., et al., Progressive left ventricular dysfunction and remodeling after myocardial infarction. Potential mechanisms and early predictors. Circulation, 1993. 87(3): p. 755–63.

32. McGhie, A.I., J.T. Willerson, and J.R. Corbett, Radionuclide assessment of ventricular function and risk stratification after myocardial infarction. Circulation, 1991. 84(3 Suppl): p. I167–76.

33. Winslow, T.M., et al., The left ventricle in systemic lupus erythematosus: Initial observations and a five-year follow-up in a university medical center population. American Heart Journal, 1993. 125(4): p. 1117–1122.

34. Nikdoust, F., et al., Early diagnosis of cardiac involvement in systemic lupus erythematosus via global longitudinal strain (GLS) by speckle tracking echocardiography. J Cardiovasc Thorac Res, 2018. 10(4): p. 231–235.

35. Taha, M., et al., Subclinical left ventricular dysfunction during systemic lupus erythematosus activity with follow-up after remission – A speckle tracking echocardiographic study. The Egyptian Rheumatologist, 2022. 44(4): p. 279–285.

36. Zhong, X., et al., Early assessment of subclinical myocardial injury in systemic lupus erythematosus by two-dimensional longitudinal layer speckle tracking imaging. Quantitative Imaging in Medicine and Surgery, 2022. 12(5): p. 2947–2960.

37. Bray, C., et al., Erythrocyte Sedimentation Rate and C-reactive Protein Measurements and Their Relevance in Clinical Medicine. WMJ, 2016. 115(6): p. 317–21.

38. Braggion Santos, M.F., et al., Myocardial late gadolinium enhancement in systemic lupus erythematosus as a marker of chronic inflammation. Journal of Cardiovascular Magnetic Resonance, 2013. 15(1): p. P121.

39. Haque, S., et al., Progression of subclinical and clinical cardiovascular disease in a UK SLE cohort: the role of classic and SLE-related factors. Lupus Sci Med, 2018. 5(1): p. e000267.

40. Roman, M.J., et al., Prevalence and correlates of accelerated atherosclerosis in systemic lupus erythematosus. N Engl J Med, 2003. 349(25): p. 2399–406.

41. Roman, M.J., et al., Rate and determinants of progression of atherosclerosis in systemic lupus erythematosus. Arthritis Rheum, 2007. 56(10): p. 3412–9.

42. Tsao, C.W., et al., Left Ventricular Structure and Risk of Cardiovascular Events: A Framingham Heart Study Cardiac Magnetic Resonance Study. J Am Heart Assoc, 2015. 4(9): p. e002188.

43. Bluemke, D.A., et al., The relationship of left ventricular mass and geometry to incident cardiovascular events: the MESA (Multi-Ethnic Study of Atherosclerosis) study. J Am Coll Cardiol, 2008. 52(25): p. 2148–55.

44. de Simone, G., et al., Left ventricular mass predicts heart failure not related to previous myocardial infarction: the Cardiovascular Health Study. Eur Heart J, 2008. 29(6): p. 741–7.

45. Mavrogeni, S., et al., Cardiovascular magnetic resonance in autoimmune rheumatic diseases: a clinical consensus document by the European Association of Cardiovascular Imaging. Eur Heart J Cardiovasc Imaging, 2022. 23(9): p. e308–e322.

